# Assessing the impact of social prescribing on health service utilisation: Evidence from the UK

**DOI:** 10.64898/2026.01.30.26345222

**Authors:** Feifei Bu, Jessica Sarah Kurland, Daniel Hayes, Daisy Fancourt

## Abstract

Social prescribing (SP) is growing rapidly in the United Kingdom and internationally. However, the evidence for its impact is still limited. Drawing pre-post data from longitudinal administrative records (n=4,547), this study aimed to investigate whether SP has the potential to reduce health service utilisation in both primary and secondary care settings. The outcomes were measured using self-reported GP visits, A&E attendances and hospital admissions in the last three months. Data were analysed using Bayesian growth curve modelling, with Poisson or hurdle lognormal models tailored to the specific outcome. Our findings demonstrate consistent patterns of reduced health service utilisation across all outcome measures and model components. Specifically, GP attendance decreased by an average of 1 visit per person (95% CI: -1.07 to -0.95) in three months following SP (53.1% reduction). A&E attendance decreased by 0.04 admissions per person (95% CI: -0.06 to -0.03), equivalent to a 62.6% reduction. And hospital admissions decreased by 0.03 admission per person (95% CI: -0.03 to -0.02), equivalent to a 61.7% reduction. We found limited evidence that the health service utilisation changes differ across sociodemographic groups, indicating a broad applicability of SP interventions.

## Introduction

The healthcare system in the United Kingdom (UK) is under intense strain, partly driven by demographic change and population ageing, and exacerbated by the COVID-19 pandemic and funding pressures. Over the past decade, the National Health Service (NHS) has experienced increasing demand, particularly from a growing prevalence of complex health problems including mental health and other long-term conditions^1^. Within primary care, an estimated 19% of general practitioner (GP) time are primarily for reasons that are non-medical^2^, but often relate to social determinants that underpin mental and physical illness. Amongst various measures being implemented to help personalise care to individual health and social needs, social prescribing (SP) has been introduced into the NHS Long Term Plan 2019^3^. SP is a mechanism of care that connects people to non-clinical support within their communities (e.g. arts, nature, exercise, volunteering, training, advice) via ‘what matters to you’ conversations to improve health and wellbeing^4,5^. The most common SP model in the UK involves a GP making a referral to a Link Worker (LW), who then collaborates with the patient, connecting them with community resources and interventions tailored to their needs and interests^5^. Although less common, individuals can also be referred via alternative routes, such as secondary care, local authorities, educational settings, voluntary, community, and social enterprise (VCSE) organisations, and self-referral. Currently, there are around 3,300 LWs employed in the NHS, and further expansion is planned as part of the newly-announced neighbourhood health service within the 2025 NHS 10-Year Plan^6^.

The rationale for developing SP was not based on trying to change patterns of healthcare utilisation amongst patients, but instead to provide a more comprehensive model of personalised care^7^. This reflects a shift from the traditional biomedical focus to a more holistic understanding of health, incorporating other complementary treatments or interventions^5^. However, questions have been raised as to whether SP has the potential to alleviate some of the pressure on the healthcare system by reducing health service utilisation^8,9^. Theoretically, the rationale for such effects is complex. First, SP has been identified as an effective way of improving health behaviours (e.g. physical activity) and certain health outcomes (e.g. mental health and wellbeing), which could ultimately reduce healthcare need and service utilisation as patients become healthier and better at managing their own health^10,11^. Moreover, for a sub-set of eligible patients who are lonely or make very frequent use of GP services, SP may provide a more appropriate forum than a GP consultation for social connection and support^12–14^. However, SP could also *increase* health service utilisation in a beneficial way. If SP can increase patient activation (which broadly refers to the knowledge, skills, ability, and motivation an individual has to manage their own health and healthcare^15^), particularly for those with low levels of activation initially, these patients may make more proactive use of health services (particularly elective or preventative services)^16^. This relationship is noted in large-scale epidemiological research, where increased engagement with community services have been related to increased primary care utilisation alongside longer-term reduced engagement with secondary and tertiary services^17^. Additionally, given SP is particularly targeted at individuals with complex social or long-term needs, it is possible that patients referred to the service may be those who are accumulating additional health needs that appropriately require increased health service utilisation. As such, SP referrals may not causally lead to increased health service utilisation but may be a symptom of increasing need. Therefore, assessing the effect of SP on health service utilisation also necessitates a person-centred lens that considers heterogeneity of effects.

In line with this complex theoretical backdrop, empirical evidence on the impacts of SP on health service utilisation is mixed. A systematic review of largely small-scale local SP evaluations found small to moderate effects in reducing GP visits and A&E attendance^18^. However, the same review also included contrasting evidence from a few studies reporting no change or an increase in service utilisation following SP^18^. Another review from the same year found little to no evidence on GP visits and insufficient evidence on hospitalisation^19^. However, most of the UK-based studies included in these reviews predated the national roll-out of SP in 2019, and were undertaken during a period characterised by pilot projects with varying infrastructure, scope and design. Therefore, research from this early phase may have limited applicability to the more established and system-wide service delivery in place today. Additionally, most of these studies were further limited by weak methodological quality, including issues of small sample size and inadequate statistical power. A more recent evidence synthesis across nine local health systems in England led by the National Academy for Social Prescribing (NASP) reported consistent and substantial reduction in GP visits, A&E attendance and hospital admissions^20^. However, most of these evaluations were not peer-reviewed or methodologically robust, employing oversimplified analytical approaches without quantifying uncertainty. Therefore, there is an urgent need for high-quality research with rigorous designs to understand the impact of SP on health service utilisation.

In response to the evidence gap, the present study drew data from Access Elemental, the most widely adopted digital SP platform in the UK. Access Elemental is a key dataset as it has a large sample size, spans varied geographical, demographic, and service contexts, and includes referrals made through both GP and non-GP SP referral routes, thereby providing a more diverse picture of SP implementation than provided through analyses solely of GP records. Using a pre-post study design, the present study aimed to examine changes in health service utilisation following SP, including GP appointments, A&E visits, and hospital admissions, and to understand whether changes vary across key sociodemographic patient characteristics.

## Methods

### Data

This study used data from Access Elemental, a digital SP platform used by health and social care professionals, community development workers and other service providers to keep track of SP activities and their impact from the point of referral. By April 2025, Access Elemental had data from over 600,000 SP cases from 482,575 patients (accessed on 25/04/2025). A detailed description of the Access Elemental data is available elsewhere^21,22^. In this study, data were restricted to patients with repeated measures of health service utilisation between two time points within 3-12 months between 2017-2025 in the UK, with a mean follow-up of 5.3 months (SD=2.0). For a small number of patients with more than one separate SP referral recorded (4%), the first case was used. This provided an analytical sample of 4,547 patients from over 120 SP sites across the UK. Among this full analytical sample, 3,651 participants (80.3%) had no missing data on covariates. See the sample selection diagrams in the Supplementary Material (Figure S1). All informed consent procedures were administered by the individual SP sites during service delivery. Access Elemental secured opt-out consent from these sites for anonymised data sharing. The research project was approved by the UCL Research Ethics Committee (23909/002).

### Measurement

Health service utilisation was measured by asking patients retrospectively at different timepoints how many times they have attended (1) general practice (GP) or (2) Accident & Emergency (A&E) in the last three months, and (3) how many times they have been admitted to a hospital in the last three months. These questions were typically administered during the first and final sessions with the link worker.

Sociodemographic covariates included gender (female, male), age (under 30, 30-49, 50-69, 70+), area deprivation (quintiles: 1-most deprived to 5-least deprived), and urbanicity (urban, rural). Both area deprivation and urbanicity were derived via postcode linkage to urban rural classification and Index of Multiple Deprivation (IMD) at Lower Layer Super Output Area (LSOA) in each UK country. We also included referral route indicating whether patients were referred from medical (e.g. GP, secondary care) or non-medical routes (e.g. education, local authority). Although ethnicity was available, as an optional measure, it had a high missing rate (>83%), so was excluded from the analysis.

### Statistical analysis

Data were analysed using Bayesian growth curve modelling. This allowed us to examine person-specific and average changes between two time points, with a random intercept and slope. For each health service utilisation outcome, we fitted an unconditional growth model to the full analytical sample (n=4,547). Further analyses were conducted to examine if changes in health service utilisation differed by individual characteristics by fitting conditional growth models using data from the subsample without missing covariates (n=3,651). Specifically, GP attendance as count data was fitted using Poisson models. Both A&E attendance and hospital admission had excessive zeros, so were fitted using hurdle lognormal models, which involve two separate processes: (1) a hurdle model (hu) predicting if the outcome is zero, and (2) lognormal model (mu) predicting the value of outcome if not zero. The Bayesian models were fitted using non-informative priors, a minimum of 4000 iterations, a burn-in of 1000, 4 Markov chains, and a thinning of 5. All analyses were conducted in in R 4.4.1 using the brms package^23^. Marginal effects were obtained using the emmeans package^24^.

## Results

As shown in Table 1, the analytical sample was broadly comparable to the full Access Elemental cohort across key sociodemographic factors (e.g. gender, age, area deprivation) and year. However, we observed an overrepresentation of individuals from rural areas and medical routes, and notably for individuals from Scotland and Northern Ireland, with very few from Wales. This pattern likely reflects differences in the preference for outcome measures across countries and SP models.

**Table 1.** Sample characteristics compared to the full Access Elemental cohort.

|  | Analytical sample |  |  | Full cohort |  |  |
| --- | --- | --- | --- | --- | --- | --- |
|  | Valid n | Freq. | % | Valid n | Freq. | % |
| Gender: female | 4,530 | 3,000 | 66.2% | 584,255 | 360,846 | 61.8% |
| Gender: male | 4,530 | 1,530 | 33.8% | 584,255 | 223,409 | 38.2% |
| Age: <30 | 4,513 | 448 | 9.9% | 597,684 | 91,021 | 15.2% |
| Age: 30-49 | 4,513 | 1,330 | 29.5% | 597,684 | 188,671 | 31.6% |
| Age: 50-69 | 4,513 | 1,792 | 39.7% | 597,684 | 190,543 | 31.9% |
| Age: 70+ | 4,513 | 943 | 20.9% | 597,684 | 127,449 | 21.3% |
| IMD 1 | 4,328 | 1,298 | 30.0% | 468,524 | 173,886 | 37.1% |
| IMD 2 | 4,328 | 971 | 22.4% | 468,524 | 104,686 | 22.3% |
| IMD 3 | 4,328 | 905 | 20.9% | 468,524 | 77,169 | 16.5% |
| IMD 4 | 4,328 | 851 | 19.7% | 468,524 | 66,585 | 14.2% |
| IMD 5 | 4,328 | 303 | 7.0% | 468,524 | 46,198 | 9.9% |
| Area: rural | 4,328 | 1,154 | 26.7% | 468,524 | 56,902 | 12.1% |
| Area: urban | 4,328 | 3,174 | 73.3% | 468,524 | 411,622 | 87.9% |
| Referral route: non-medical | 3,830 | 215 | 5.6% | 542,663 | 92,546 | 17.1% |
| Referral route: medical | 3,830 | 3,615 | 94.4% | 542,663 | 450,117 | 82.9% |
| Country: England | 4,547 | 1,919 | 42.2% | 600,001 | 495,075 | 82.5% |
| Country: Wales | 4,547 | 4 | 0.1% | 600,001 | 60,620 | 10.1% |
| Country: Scotland | 4,547 | 1,251 | 27.5% | 600,001 | 16,141 | 2.7% |
| Country: Northern Ireland | 4,547 | 1,373 | 30.2% | 600,001 | 28,165 | 4.7% |
| Case year: 2019 | 4,547 | 485 | 10.7% | 601,529 | 22,824 | 3.8% |
| Case year: 2020 | 4,547 | 399 | 8.8% | 601,529 | 45,241 | 7.5% |
| Case year: 2021 | 4,547 | 881 | 19.4% | 601,529 | 85,719 | 14.3% |
| Case year: 2022 | 4,547 | 1,048 | 23.0% | 601,529 | 126,991 | 21.1% |
| Case year: 2023 | 4,547 | 748 | 16.5% | 601,529 | 136,863 | 22.8% |
| Case year: 2024 | 4,547 | 976 | 21.5% | 601,529 | 128,472 | 21.4% |
| Case year: 2025 | 4,547 | 10 | 0.2% | 601,529 | 43,373 | 7.2% |

Figure 1 presents the expected number of GP attendances per person and their 95% credible intervals (CI) from posterior distributions of unconditional growth models (also see Table S1). GP attendance decreased by an average of 1 visit per person (95% CI: -1.07 to -0.95) between the pre- and post-intervention periods over an outcome assessment window of 3 months (Figure 1). This was equivalent to a 53.1% reduction in the number of GP attendance, from 1.89 (95% CI: 1.84 to 1.96) to 0.89 (95% CI: 0.85 to 0.93). The random estimates showed substantial individual heterogeneities in both the intercept and slope (Table S1). And the weak positive correlation between them (0.38, 95% CI: 0.25 to 0.52) suggested patients with higher number of GP attendance had smaller reductions.

**Figure 1.**
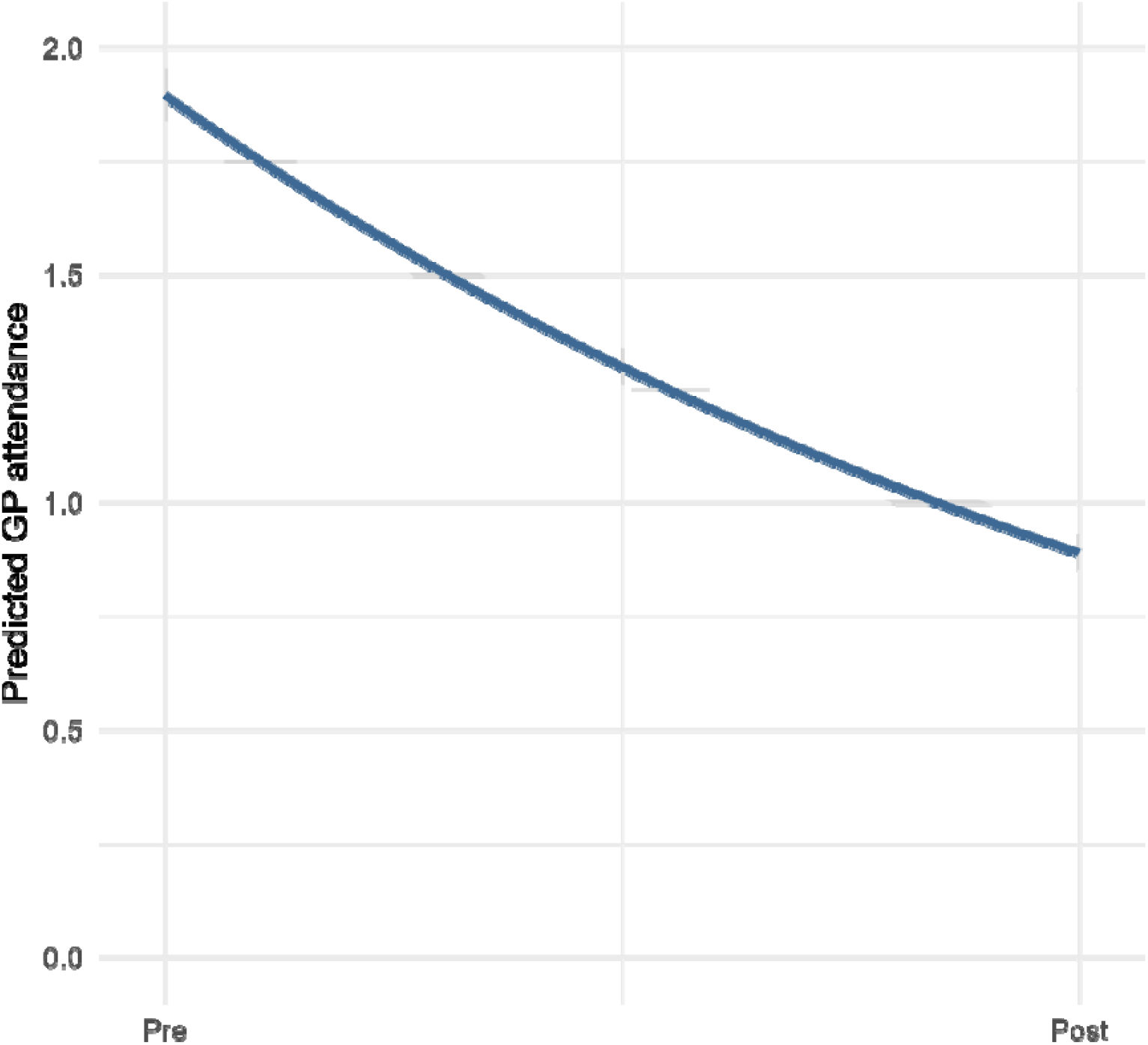
Predicted GP attendance between pre- and post-intervention periods based on unconditional growth model with Poisson family

Results integrating the hurdle and lognormal models showed that the number of A&E attendances per person decreased from 0.07 (95% CI: 0.06 to 0.09) to 0.03 (95% CI: 0.02 to 0.03) between pre- and post-intervention periods. This represents an absolute reduction of 0.04 admissions per person (95% CI: -0.06 to -0.03), equivalent to a 62.6% decrease (Figure 2a). Based on the hurdle model (Figure 2b), the probability of having no A&E attendance increased from 94.4% (95% CI: 93.3% to 95.5%) to 97.8% (95% CI: 97.2% to 98.4%). Focusing on individuals with non-zero attendance, the number of A&E attendances per person decreased from 1.23 (95% CI: 1.19 to 1.27) to 1.16 (95% CI: 1.11 to 1.21), representing an absolute reduction of 0.06 attendance (95% CI: -0.12 to -0.01), equivalent to 5.1% decrease (Figure 2c; full results are presented in Supplementary Table S2).

**Figure 2.**
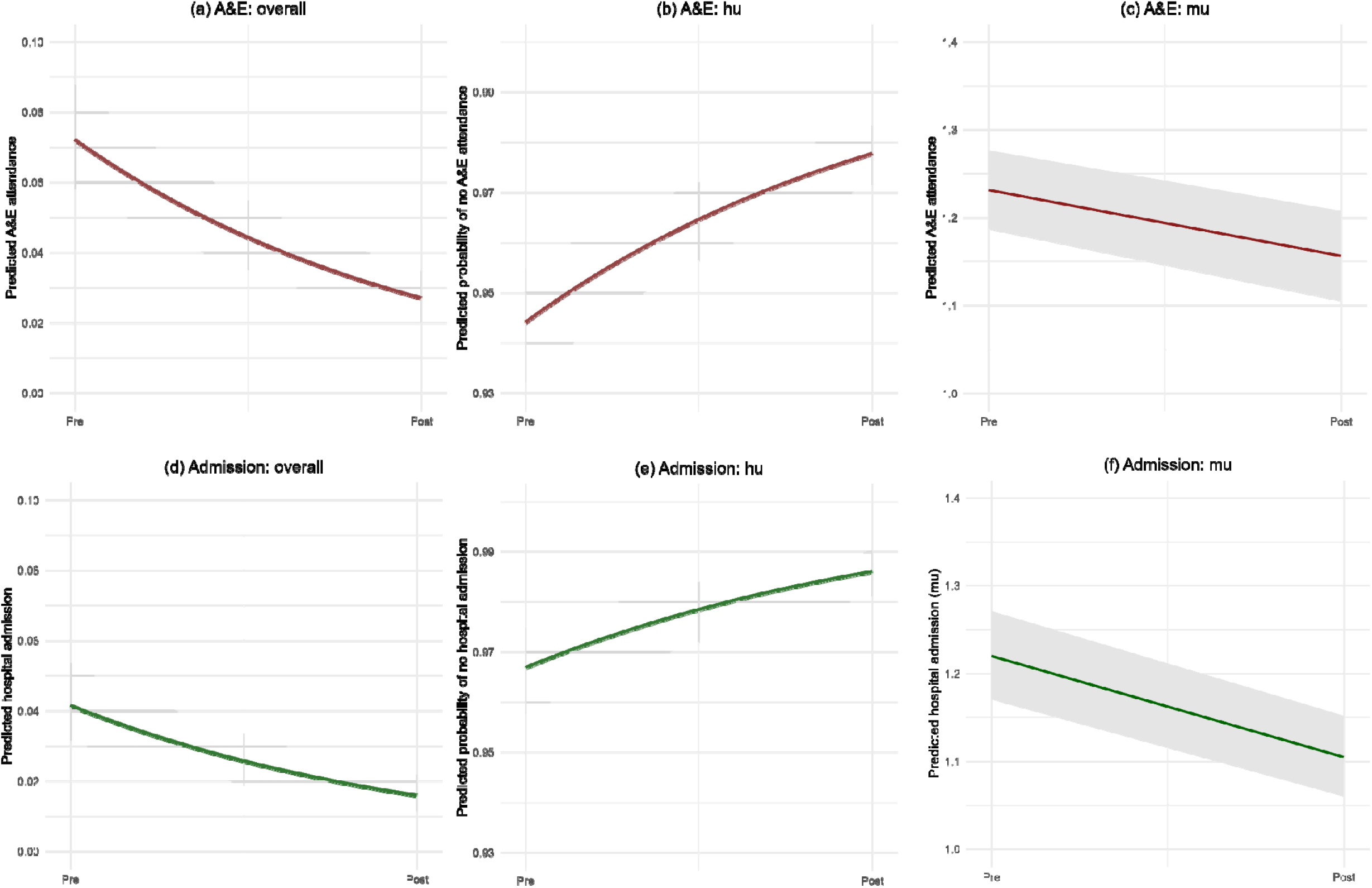
Predicted A&E attendance and hospital admissions between pre- and post-intervention periods based on unconditional growth model with hurdle lognormal family Reference characteristics: (a-b) female, age 50-69, first IMD quintile, from medical route (c) female, age 50-69, first IMD quintile, urban area (d-e) female, first IMD quintile, urban, from medical route

Overall, the hospital admission number per person decreased from 0.04 (95% CI: 0.03 to 0.05) to 0.02 (95% CI: 0.01 to 0.02) between pre- and post-intervention periods, representing an absolute reduction of 0.03 admission per person (95% CI: - 0.03 to -0.02), equivalent to a 61.7% decrease (Figure 2d). Specifically, the hurdle model (Figure 2e) revealed that the probability of having no hospital admission increased from 96.7% (95% CI: 95.8% to 97.5%) to 98.6% (95% CI: 98.1% to 99.1%). Focusing on individuals with non-zero admission, the number of hospital admissions per person decreased from 1.22 (95% CI: 1.17 to 1.27) to 1.10 (95% CI: 1.06 to 1.15). This represents an absolute reduction of 0.1 (95% CI: -0.15 to -0.05) admissions, corresponding to an 8.2% decrease (Figure 2e; full results are presented in Supplementary Table S3). Notably, for both A&E and hospital admission, there was a strong negative correlation between the intercept and slope (-0.72 and - 0.81 respectively), suggesting that patients starting with higher attendance or admission had greater reductions.

Results from the conditional models showed limited evidence that changes in health service utilisation differed by individual characteristics, with a few exceptions (Table S4-S6). For GP attendance, the decrease measured by incident rate ratios (IRR) was greater among people living in rural areas (IRR: 0.34, 95% CI: 0.25 to 0.45) than their urban counterparts (IRR: 0.47, 95% CI: 0.35 to 0.62). However, the difference was largely negligible when converted back to counts (Figure 3a). For A&E attendance, changes in the probability of any attendance were also greater for rural residents (Figure 3b). Further, there was little change in A&E attendance probability amongst individuals from non-medical routes, while for those referred to SP through medical routes, the probability of having no A&E attendance increased from 92.2% (95% CI: 89.6% to 94.8%) to 97.6% (95% CI: 96.4% to 98.7%; Figure 3c). There was some evidence that the changes in hospital admissions differed by age groups. Specifically, older adults aged 70 or older experienced little change. For people under 30, the decreases between the pre- and post-intervention periods occurred in the number of admissions, while older age groups (aged 30-69) experienced decreases in the probability of any admission (Figure 3d and 3e).

**Figure 3.**
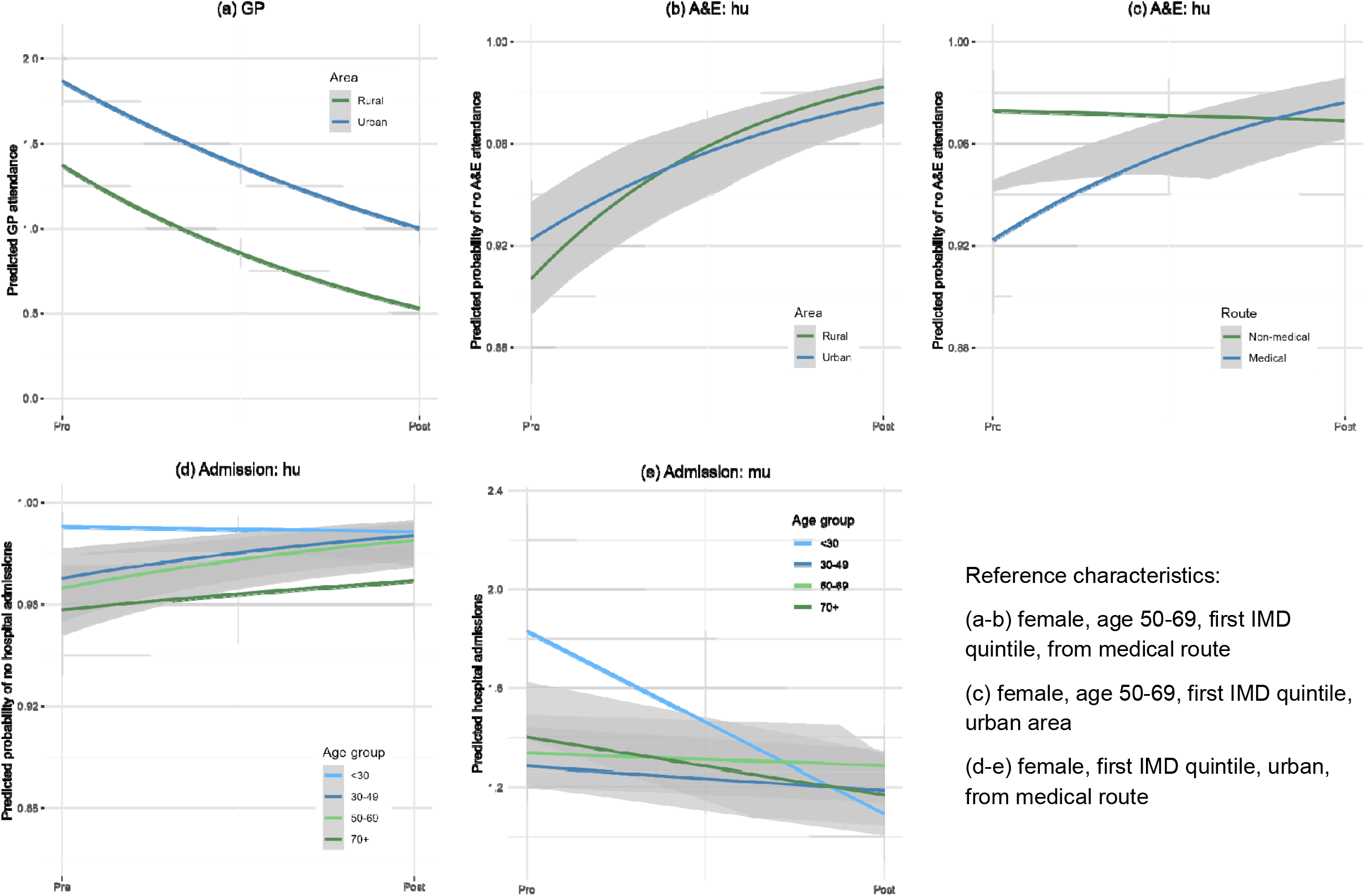
Predicted healthcare service utilisation between pre- and post-intervention periods by individual characteristics based on conditional growth models

## Discussion

Analysing data from 4,547 patients from over 120 SP sites across the UK, our study is one of the first large-scale national analyses of SP’s impact on health service utilisation. Our findings provide robust empirical evidence suggesting SP may relate to reductions in both primary and secondary health service utilisation, including GP visits, A&E attendance and hospital admissions. There was some but limited evidence of differing effects across sociodemographic groups.

Our findings demonstrate a consistent pattern of reduced health service utilisation across all outcome measures and model components following SP programmes. On average, SP is associated with one fewer GP visit per patient in a three-month outcome assessment window, equivalent to a 53.1% reduction. This estimate is largely comparable to findings from local evaluations (with the same follow-up period) in Tameside and Glossop (49.5%), as well as in Kirklees focusing on frequent service users (50%)^20^. For A&E attendance and hospital admissions, our analyses indicated decreases in both the probability of using these services and the frequency of utilisation among patients with at least one attendance or admission, relating to overall decreases of 62.6% for A&E attendance and 61.7% for hospital admission. Whilst these reductions are small in absolute magnitude, due to the low frequency of secondary care use at baseline, they could nonetheless translate into substantial cost savings due to the high unit costs of such services^25^ and the large number of SP referral made annually.

Consistent with previous work on wellbeing outcomes^26^, we found limited evidence that the changes in health service utilisation following SP differ by individual characteristics. Slight variations in GP attendance based on urbanicity were statistically significant but materially almost negligible. Although there was some evidence that changes in probability of A&E attendance differ by urbanicity and referral route, this is likely driven by differences in baseline probability. For instance, people from non-medical routes start from a lower probability of A&E attendance (likely driven by distinct patient health profiles across medical and non-medical referrals), leaving limited scope for a measurable reduction (i.e. a floor effect). For hospital admissions, the only factor moderating the rate of change was age, with limited change in the probability of hospital admission for patients under 30 or aged 70 or older. For the younger age group, this likely again reflects a floor effect, as baseline probability is already very low, while for older adults aged 70 or older, preventing admissions is inherently more challenging due to the presence and potential complexity of long-term conditions^27,28^. Interestingly, however, the decrease in frequency for patients with non-zero admission was most prominent for patients under 30. This suggests that for younger adults who are hospitalised, SP may have a particularly preventative effect; a finding that merits further investigation.

Overall, when explored against the complex question of whether SP impacts health service utilisation, there is support that SP may reduce health service use across GP appointments, A&E attendance, as well as hospital admissions. This effect may be driven by SP having a positive impact on patients’ health behaviour, and overall health and wellbeing, resulting in reduced need for healthcare services. In particular, previous evidence suggests SP may be beneficial for mental health and wellbeing^11^, which represent the most common reasons for SP referral^29^. A recent qualitative paper exploring SP for youth with mental health difficulties lends support to this showing that one mechanism that SP activates is better emotional recognition and regulation^30^. Another potential mechanism is through enhanced social connection^14^. SP is uniquely positioned to provide social capital and social support (e.g. emotional, practical, informational) either directly through structured link workers sessions and referral to community resources, or indirectly by fostering expanded social networks, which consequently may contribute to improved health self-management and reduced health service utilisation^31^. It is important to note that while our study supports an average decrease in healthcare utilisation following SP, individual trajectories can be highly heterogeneous. This is in line with theory that suggests SP may *increase* healthcare utilisation through increased patient activation and subsequent increased proactive use of healthcare services^16^, with some studies finding increases or no change^18^. In addition to individual heterogeneity, the divergent intercept-slope correlations for primary and secondary care are also worth noting--positive for GP attendance and negative for A&E and hospital admissions. This suggests that individuals with higher baseline number of GP attendances experienced smaller reductions, whereas those with more A&E attendance or hospital admissions experienced greater reductions. This suggests that the mechanism by which SP influences service utilisation may differ between primary and secondary care settings, warranting further investigation.

A primary methodological strength of our study is analysing routinely collected data from a large-scale cloud-based platform that enables tracking of diverse SP referral pathways across more than 120 SP sites in the UK. Further, we have employed robust analytical models that account for heterogeneous treatment effects across patients and the correlation of random effects. However, the present study has several important limitations. First, our analyses are restricted to a subsample of the Access Elemental cohort with pre-post health service utilisation measures. While the analytical sample is shown to have largely comparable sociodemographic profiles to the whole cohort, unbalances were found in countries, areas and referral routes. We also cannot rule out the possibility of selection bias related to unmeasured characteristics, or due to the overall representativeness of the Access Elemental cohort. Second, as self-reported measures, health service utilisation may be subject to recall bias or social desirability. Future effort is needed to link SP data to administrative health records to eliminate such biases (although it is noted that analyses of administrative records bring their own biases too, such as complexities and discrepancies in coding practices). Third, the absence of a control group presents a major constraint. Uncontrolled pre-post designs are vulnerable to regression to the mean; a statistical artifact that occurs when participants have extreme baseline values, leading to a natural decline when retested even without any intervention^32^. However, our analyses account for baseline-dependent growth by allowing for correlations between random intercepts and slopes and also allow us to inspect person specific changes, in addition to the average rate of change, which mitigates some underlying causes of regression to the mean. Finally, we focus on the period from 3-12 months following SP referral (looking consistently at the 3-month period prior to follow-up). Thus, our analyses do not provide a cumulative understanding of how SP influences health service utilisation over timescales beyond 3 months, nor explore other types of health service utilisation (e.g. nursing home, dental or outpatient care). We particularly recommend the need for long-term tracking of health service utilisation amongst patients referred to SP.

## Conclusion

Our study advances the evidence base for SP by demonstrating consistent reductions in health service utilisation across different measures, using national data and robust analytical approaches. Nevertheless, further evidence from high-quality observational or controlled studies remains essential to consolidate the effectiveness of SP, understand its mechanisms of action, and identify which components work best for specific populations. Only through such rigorous and ongoing inquiry can the potential of SP be fully understood and optimally integrated into the evolving landscape of neighbourhood health model.

## Acknowledgements

This work was supported by the Economic and Social Research Council (ESRC) [UKRI1717], Arts and Humanities Research Council (AHRC) [AH/W006405/1] and the National Academy for Social Prescribing (NASP).

## Data availability statement

The data used in this study are subject to third-party restrictions, so are not public available. Data are however available from the authors upon reasonable request, subject to permission from the Access Elemental.

