## Supplement for "Assessing the impact of social prescribing on health service utilisation: Evidence from the UK"

### Supplementary Materials

Table S1 Results from the Bayesian unconditional growth curve model with Poisson family on GP attendance

| Parameter | Coef. | 95% CI | Rhat | Bulk_ESS | Tail_ESS |
|---|---|---|---|---|---|
| Intercept | 0.64 | [0.61, 0.67] | 1.00 | 2377 | 2413 |
| wave | -0.76 | [-0.8, -0.71] | 1.00 | 2314 | 2143 |
| SD (Intercept) | 0.69 | [0.66, 0.71] | 1.00 | 2228 | 1883 |
| SD (wave) | 0.38 | [0.32, 0.43] | 1.00 | 1407 | 2058 |
| Cor (Intercept, wave) | 0.38 | [0.25, 0.52] | 1.00 | 1477 | 2139 |

Table S2. Results from the Bayesian unconditional growth curve model with hurdle lognormal family on A&E attendance

| Parameter | Coef. | 95% CI | Rhat | Bulk_ESS | Tail_ESS |
|---|---|---|---|---|---|
| Intercept | 0.21 | [0.17, 0.24] | 1.00 | 3360 | 3262 |
| wave | -0.06 | [-0.12, -0.01] | 1.00 | 3616 | 3772 |
| hu: Intercept | 2.83 | [2.62, 3.05] | 1.00 | 2668 | 3601 |
| hu: wave | 0.96 | [0.78, 1.15] | 1.00 | 4049 | 3967 |
| SD (Intercept) | 0.28 | [0.19, 0.38] | 1.01 | 201† | 222† |
| SD (wave) | 0.20 | [0.03, 0.41] | 1.01 | 202† | 241† |
| SD (hu: Intercept)‡ | 1.57 | [1.34, 1.80] | 1.00 | 2539 | 3367 |
| Cor (Intercept, wave) | -0.72 | [-0.98, -0.34] | 1.00 | 1941 | 2200 |
| Sigma (residual) | 0.30 | [0.78, 1.15] | 1.00 | 192† | 140† |

Notes: † low ESS, the specific results should be interpreted with caution, ‡ hurdle model fitted with random intercept only for model convergence

Table S3. Results from the Bayesian unconditional growth curve model with hurdle lognormal family on hospital admissions

| Parameter | Coef. | 95% CI | Rhat | Bulk_ESS | Tail_ESS |
|---|---|---|---|---|---|
| Intercept | 0.20 | [0.16, 0.24] | 1.00 | 3871 | 3806 |
| wave | -0.10 | [-0.15, -0.04] | 1.00 | 3788 | 3892 |
| hu: Intercept | 3.37 | [3.11, 3.65] | 1.00 | 3255 | 3468 |
| hu: wave | 0.89 | [0.69, 1.09] | 1.00 | 3995 | 3893 |
| SD (Intercept) | 0.37 | [0.3, 0.44] | 1.05 | 75† | 136† |
| SD (wave) | 0.28 | [0.14, 0.43] | 1.05 | 74† | 164† |
| SD (hu: Intercept)‡ | 1.84 | [1.6, 2.11] | 1.00 | 2976 | 3713 |
| Cor (Intercept, wave) | -0.81 | [-0.99, -0.67] | 1.03 | 136† | 1261 |
| Sigma (residual) | 0.21 | [0.07, 0.30] | 1.07* | 59† | 60† |

Notes: * Rhat>1.05, † low ESS, the specific results should be interpreted with caution, ‡ hurdle model fitted with random intercept only for model convergence

Table S4 Results from the Bayesian conditional growth curve model with Poisson family on GP attendance

| Parameter | Coef. | 95% CI | Rhat | Bulk_ESS | Tail_ESS |
|---|---|---|---|---|---|
| Intercept | -0.66 | [-0.87, -0.45] | 1.00 | 3722 | 3714 |
| Wave | -1.09 | [-1.38, -0.80] | 1.00 | 3686 | 3714 |
| Female | 0.11 | [0.04, 0.18] | 1.00 | 4048 | 3659 |
| Age 30-49 | 0.32 | [0.20, 0.45] | 1.00 | 4101 | 4003 |
| Age 50-69 | 0.26 | [0.14, 0.38] | 1.00 | 3806 | 3875 |
| Age 70+ | 0.16 | [0.03, 0.29] | 1.00 | 4074 | 4097 |
| IMD 2 | 0.16 | [0.06, 0.24] | 1.00 | 4004 | 3971 |
| IMD 3 | 0.24 | [0.15, 0.33] | 1.00 | 3830 | 3796 |
| IMD 4 | 0.15 | [0.06, 0.25] | 1.00 | 3563 | 3668 |
| IMD 5 | 0.32 | [0.18, 0.44] | 1.00 | 3944 | 3702 |
| Urban | 0.31 | [0.23, 0.39] | 1.00 | 4130 | 3904 |
| Medical route | 0.60 | [0.44, 0.77] | 1.00 | 3627 | 3972 |
| Wave: female | 0.01 | [-0.08, 0.09] | 1.00 | 3975 | 4009 |
| Wave: age 30-49 | -0.07 | [-0.22, 0.08] | 1.00 | 3919 | 3798 |
| Wave: age 50-69 | -0.02 | [-0.17, 0.14] | 1.00 | 4059 | 3859 |
| Wave: age 70+ | 0.06 | [-0.10, 0.23] | 1.00 | 3962 | 3908 |
| Wave: IMD 2 | -0.06 | [-0.18, 0.05] | 1.00 | 4203 | 3971 |
| Wave: IMD 3 | -0.09 | [-0.21, 0.02] | 1.00 | 3984 | 3798 |
| Wave: IMD 4 | -0.10 | [-0.23, 0.02] | 1.00 | 4192 | 3853 |
| Wave: IMD 5 | 0.00 | [-0.15, 0.16] | 1.00 | 3899 | 3803 |
| Wave: urban | 0.33 | [0.23, 0.43] | 1.00 | 4258 | 3968 |
| Wave: medical | 0.15 | [-0.09, 0.38] | 1.00 | 3785 | 3947 |
| SD (Intercept) | 0.67 | [0.64, 0.70] | 1.00 | 3817 | 3776 |
| SD (wave) | 0.36 | [0.29, 0.42] | 1.00 | 2454 | 3259 |
| Cor (Intercept, wave) | 0.34 | [0.18, 0.52] | 1.00 | 2772 | 3185 |

Table S5 Results from the Bayesian conditional growth curve model with hurdle lognormal family on A&E attendance

| Parameter | Coef. | 95% CI | Rhat | Bulk_ESS | Tail_ESS |
|---|---|---|---|---|---|
| Intercept | 0.05 | [-0.25, 0.35] | 1.00 | 5470 | 5479 |
| Wave | -0.10 | [-0.53, 0.32] | 1.00 | 5180 | 5455 |
| Female | 0.00 | [-0.09, 0.08] | 1.00 | 5297 | 5745 |
| Age 30-49 | 0.01 | [-0.14, 0.16] | 1.00 | 5412 | 4951 |
| Age 50-69 | 0.06 | [-0.09, 0.20] | 1.00 | 5007 | 5213 |
| Age 70+ | 0.01 | [-0.15, 0.18] | 1.00 | 5315 | 5060 |
| IMD 2 | 0.00 | [-0.11, 0.10] | 1.00 | 5493 | 5571 |
| IMD 3 | -0.04 | [-0.15, 0.08] | 1.00 | 5323 | 5471 |
| IMD 4 | 0.01 | [-0.11, 0.12] | 1.00 | 4940 | 5448 |
| IMD 5 | -0.17 | [-0.38, 0.03] | 1.00 | 5588 | 5030 |
| Urban | 0.12 | [0.02, 0.20] | 1.00 | 5878 | 5551 |
| Medical route | 0.05 | [-0.21, 0.30] | 1.00 | 5495 | 5350 |
| Wave: female | 0.02 | [-0.11, 0.15] | 1.00 | 5508 | 5613 |
| Wave: age 30-49 | -0.11 | [-0.35, 0.13] | 1.00 | 5395 | 5225 |
| Wave: age 50-69 | -0.16 | [-0.41, 0.08] | 1.00 | 5182 | 5119 |
| Wave: age 70+ | -0.10 | [-0.37, 0.15] | 1.00 | 5232 | 5318 |
| Wave: IMD 2 | 0.07 | [-0.10, 0.24] | 1.00 | 5195 | 5573 |
| Wave: IMD 3 | 0.10 | [-0.08, 0.28] | 1.00 | 5286 | 5322 |
| Wave: IMD 4 | 0.14 | [-0.05, 0.32] | 1.00 | 5088 | 5475 |
| Wave: IMD 5 | 0.10 | [-0.19, 0.40] | 1.00 | 5452 | 5340 |
| Wave: urban | -0.02 | [-0.16, 0.13] | 1.00 | 5876 | 5687 |
| Wave: medical | 0.08 | [-0.25, 0.41] | 1.00 | 5278 | 5185 |
| hu_Intercept | 3.62 | [2.70, 4.63] | 1.00 | 4794 | 5389 |
| hu_wave | 0.42 | [-0.86, 1.72] | 1.00 | 5069 | 5202 |
| hu_female | 0.10 | [-0.19, 0.37] | 1.00 | 5468 | 5384 |
| hu_age 30-49 | -0.25 | [-0.76, 0.23] | 1.00 | 5422 | 5235 |
| hu_age 50-69 | -0.33 | [-0.82, 0.13] | 1.00 | 5354 | 5408 |
| hu_age 70+ | -0.18 | [-0.71, 0.36] | 1.00 | 5598 | 5507 |
| hu_IMD_2 | 0.36 | [0.00, 0.73] | 1.00 | 5064 | 5373 |
| hu_IMD_3 | 0.53 | [0.17, 0.91] | 1.00 | 5702 | 5508 |
| hu_IMD_4 | 0.52 | [0.14, 0.92] | 1.00 | 5459 | 5515 |
| hu_IMD_5 | 1.04 | [0.41, 1.72] | 1.00 | 5710 | 4966 |
| hu_urban | 0.20 | [-0.11, 0.51] | 1.00 | 5734 | 5214 |
| hu_medical | -1.11 | [-1.94, -0.39] | 1.00 | 5386 | 4786 |
| hu_wave: female | -0.08 | [-0.52, 0.35] | 1.00 | 5371 | 5114 |
| hu_wave: age 30-49 | 0.09 | [-0.72, 0.86] | 1.00 | 5417 | 5148 |
| hu_wave: age 50-69 | 0.03 | [-0.75, 0.79] | 1.00 | 5484 | 5395 |
| hu_wave: age 70+ | -0.27 | [-1.12, 0.53] | 1.00 | 5473 | 5355 |
| hu_wave: IMD 2 | -0.02 | [-0.59, 0.54] | 1.00 | 5279 | 5607 |
| hu_wave: IMD 3 | -0.40 | [-0.97, 0.16] | 1.00 | 5065 | 5379 |
| hu_wave: IMD 4 | -0.31 | [-0.89, 0.27] | 1.00 | 5280 | 5555 |
| hu_wave: IMD 5 | -0.73 | [-1.68, 0.21] | 1.00 | 5684 | 5480 |
| hu_wave: urban | -0.50 | [-1.01, 0.00] | 1.00 | 5679 | 5139 |
| hu_wave: medical | 1.37 | [0.40, 2.37] | 1.00 | 5198 | 5255 |
| SD (Intercept) | 0.31 | [0.22, 0.40] | 1.02 | 130† | 180† |
| SD (wave) | 0.21 | [0.04, 0.41] | 1.02 | 130† | 154† |
| SD (hu_Intercept) | 1.57 | [1.30, 1.85] | 1.00 | 3580 | 4640 |
| Cor (Intercept, wave) | -0.71 | [-0.98, -0.38] | 1.00 | 2110 | 3304 |

Notes: † low ESS, the specific results should be interpreted with caution

Table S6 Results from the Bayesian conditional growth curve model with hurdle lognormal family on hospital admission

| Parameter | Coef. | 95% CI | Rhat | Bulk_ESS | Tail_ESS |
|---|---|---|---|---|---|
| Intercept | 0.56 | [-0.01, 1.13] | 1.00 | 5055 | 5099 |
| Wave | -0.36 | [-1.05, 0.32] | 1.00 | 5299 | 5294 |
| Female | 0.01 | [-0.10, 0.11] | 1.00 | 5187 | 5001 |
| Age 30-49 | -0.36 | [-0.62, -0.09] | 1.00 | 4765 | 5102 |
| Age 50-69 | -0.32 | [-0.57, -0.06] | 1.00 | 4944 | 5180 |
| Age 70+ | -0.27 | [-0.53, 0.00] | 1.00 | 4897 | 5369 |
| IMD 2 | -0.06 | [-0.20, 0.08] | 1.00 | 5147 | 5408 |
| IMD 3 | -0.18 | [-0.33, -0.03] | 1.00 | 5144 | 5411 |
| IMD 4 | -0.06 | [-0.20, 0.09] | 1.00 | 5329 | 5123 |
| IMD 5 | -0.26 | [-0.49, -0.04] | 1.00 | 5182 | 5169 |
| Urban | 0.10 | [-0.02, 0.22] | 1.00 | 5358 | 5292 |
| Medical route | -0.05 | [-0.55, 0.44] | 1.00 | 5148 | 5340 |
| Wave: female | 0.06 | [-0.09, 0.21] | 1.00 | 5653 | 4865 |
| Wave: age 30-49 | 0.44 | [0.11, 0.76] | 1.00 | 4843 | 5407 |
| Wave: age 50-69 | 0.48 | [0.16, 0.79] | 1.00 | 4929 | 5333 |
| Wave: age 70+ | 0.33 | [0.00, 0.65] | 1.00 | 4984 | 5275 |
| Wave: IMD 2 | 0.03 | [-0.18, 0.24] | 1.00 | 5118 | 5439 |
| Wave: IMD 3 | 0.17 | [-0.04, 0.37] | 1.00 | 5120 | 5350 |
| Wave: IMD 4 | -0.02 | [-0.22, 0.18] | 1.00 | 5440 | 5487 |
| Wave: IMD 5 | 0.15 | [-0.14, 0.46] | 1.00 | 5202 | 5295 |
| Wave: urban | -0.02 | [-0.18, 0.15] | 1.00 | 5367 | 5515 |
| Wave: medical | -0.19 | [-0.77, 0.40] | 1.00 | 5411 | 5066 |
| hu_Intercept | 6.94 | [5.39, 8.74] | 1.00 | 4758 | 5278 |
| hu_wave | -1.77 | [-3.79, 0.17] | 1.00 | 5010 | 5477 |
| hu_female | 0.01 | [-0.33, 0.34] | 1.00 | 5811 | 5565 |
| hu_age 30-49 | -1.14 | [-1.96, -0.38] | 1.00 | 5069 | 5258 |
| hu_age 50-69 | -1.26 | [-2.07, -0.54] | 1.00 | 5087 | 5093 |
| hu_age 70+ | -1.50 | [-2.33, -0.73] | 1.00 | 4947 | 5193 |
| hu_IMD_2 | 0.07 | [-0.37, 0.52] | 1.00 | 5536 | 5450 |
| hu_IMD_3 | 0.25 | [-0.21, 0.72] | 1.00 | 5620 | 5408 |
| hu_IMD_4 | 0.06 | [-0.40, 0.51] | 1.00 | 5276 | 5207 |
| hu_IMD_5 | 0.38 | [-0.29, 1.08] | 1.00 | 5176 | 5377 |
| hu_urban | -0.04 | [-0.42, 0.33] | 1.00 | 5740 | 5489 |
| hu_medical | -2.28 | [-3.82, -1.05] | 1.00 | 5076 | 5075 |
| hu_wave: female | 0.21 | [-0.25, 0.68] | 1.00 | 5801 | 5147 |
| hu_wave: age 30-49 | 1.01 | [0.00, 2.04] | 1.00 | 4954 | 5356 |
| hu_wave: age 50-69 | 1.01 | [0.02, 2.02] | 1.00 | 5116 | 5409 |
| hu_wave: age 70+ | 0.51 | [-0.50, 1.51] | 1.00 | 4995 | 5213 |
| hu_wave: IMD 2 | 0.86 | [0.20, 1.55] | 1.00 | 5460 | 5625 |
| hu_wave: IMD 3 | 0.05 | [-0.58, 0.70] | 1.00 | 5425 | 5340 |
| hu_wave: IMD 4 | 0.18 | [-0.46, 0.82] | 1.00 | 5430 | 5450 |
| hu_wave: IMD 5 | 0.08 | [-0.85, 1.06] | 1.00 | 5472 | 5304 |
| hu_wave: urban | 0.01 | [-0.54, 0.54] | 1.00 | 5115 | 4850 |
| hu_wave: medical | 1.37 | [-0.23, 3.11] | 1.00 | 5390 | 5327 |
| SD (Intercept) | 0.36 | [0.24, 0.44] | 1.04 | 82† | 337† |
| SD (wave) | 0.33 | [0.10, 0.52] | 1.03 | 100† | 824† |
| SD (hu_Intercept) | 1.82 | [1.51, 2.13] | 1.00 | 3600 | 5076 |
| Cor (Intercept, wave) | -0.77 | [-0.98, -0.58] | 1.01 | 360† | 1983 |

Notes: † low ESS, the specific results should be interpreted with caution

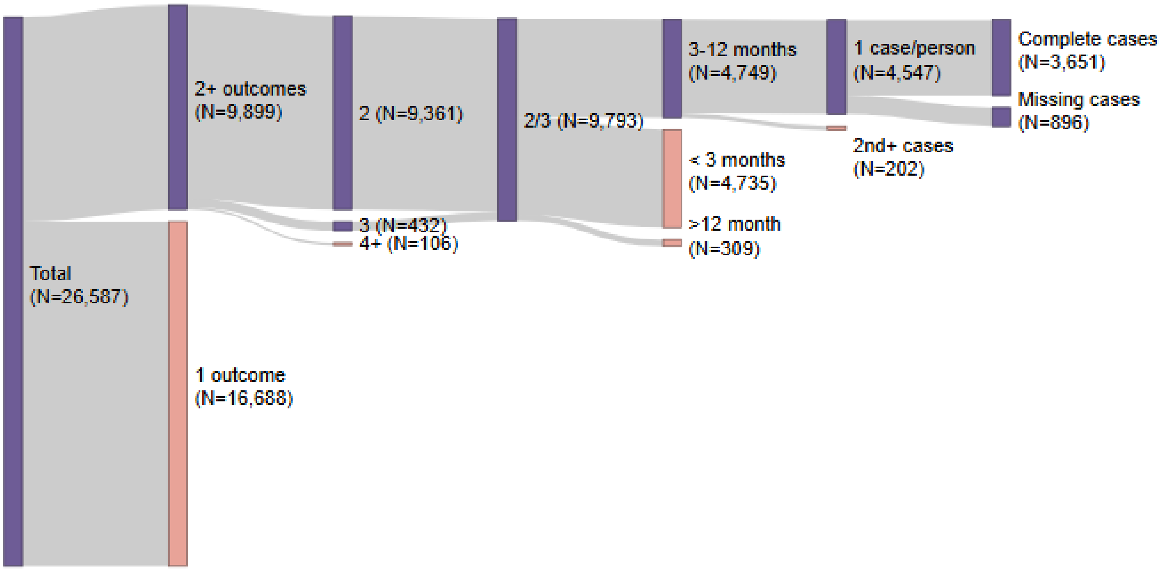

Figure S1. Analytical sample selection diagram

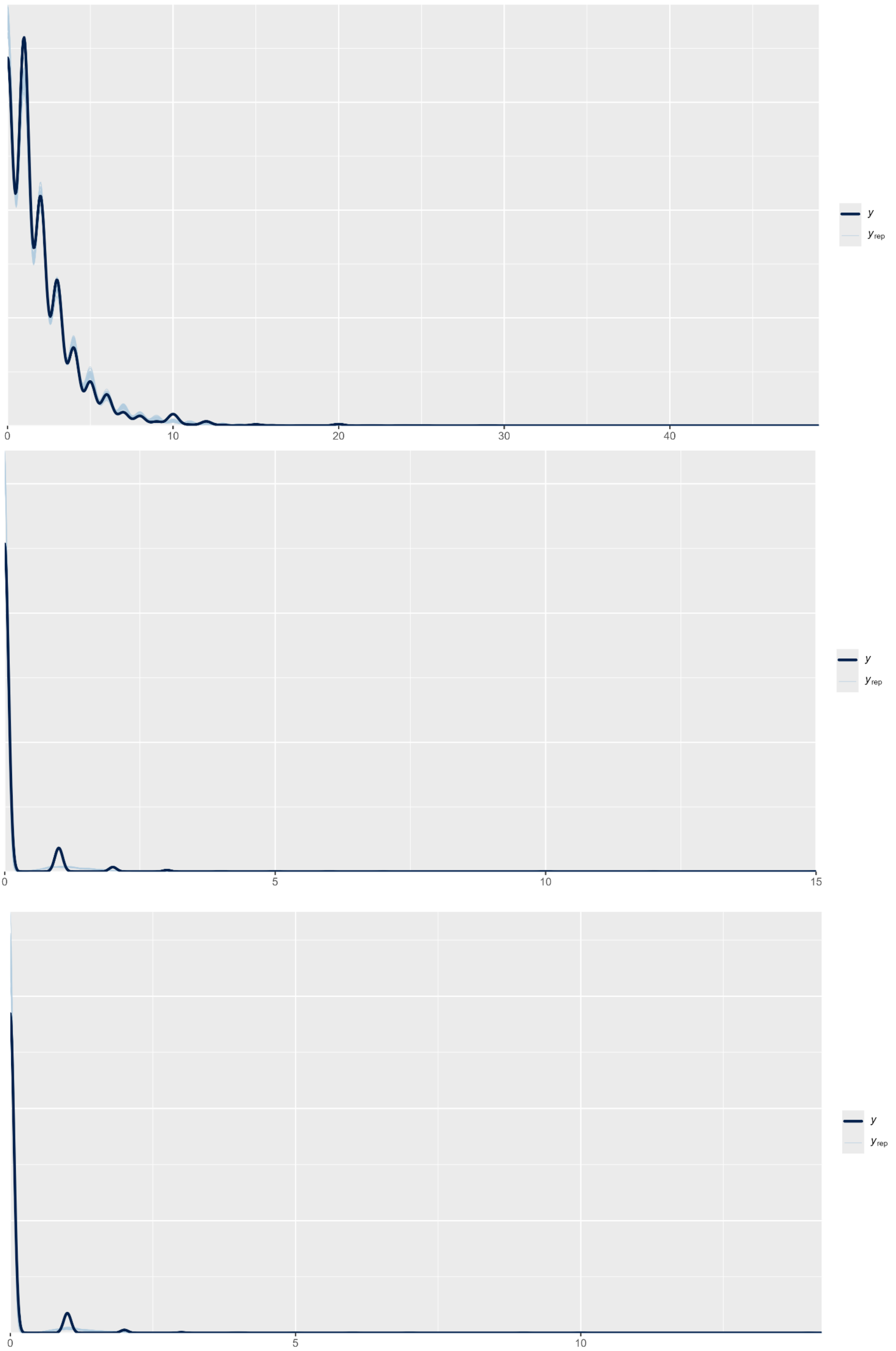

Figure S2. Posterior predictive checks for GP attendance (top), A&E attendance (middle) and hospital admission (bottom)

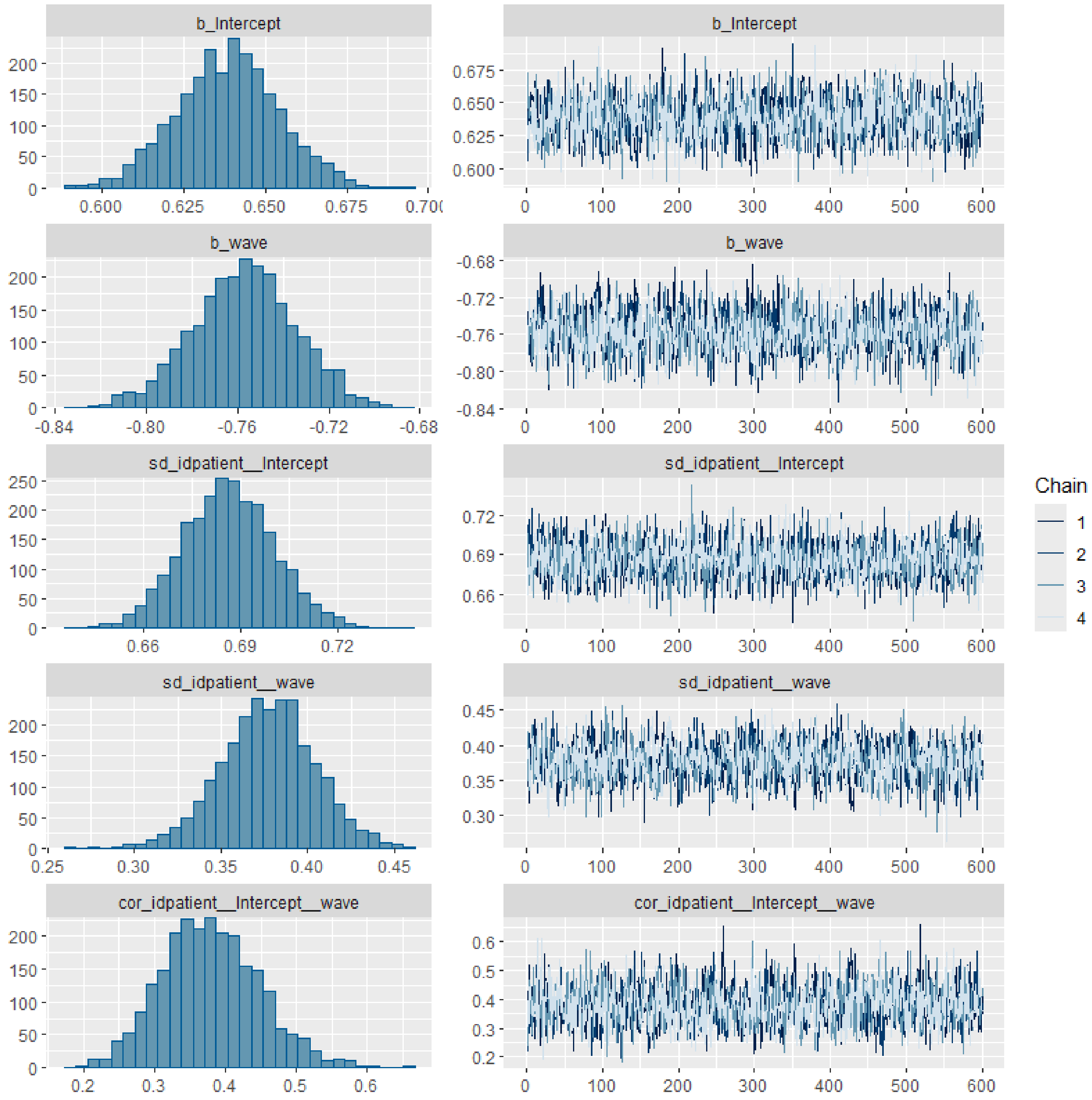

Figure S3. Trace plots for the unconditional GP attendance model

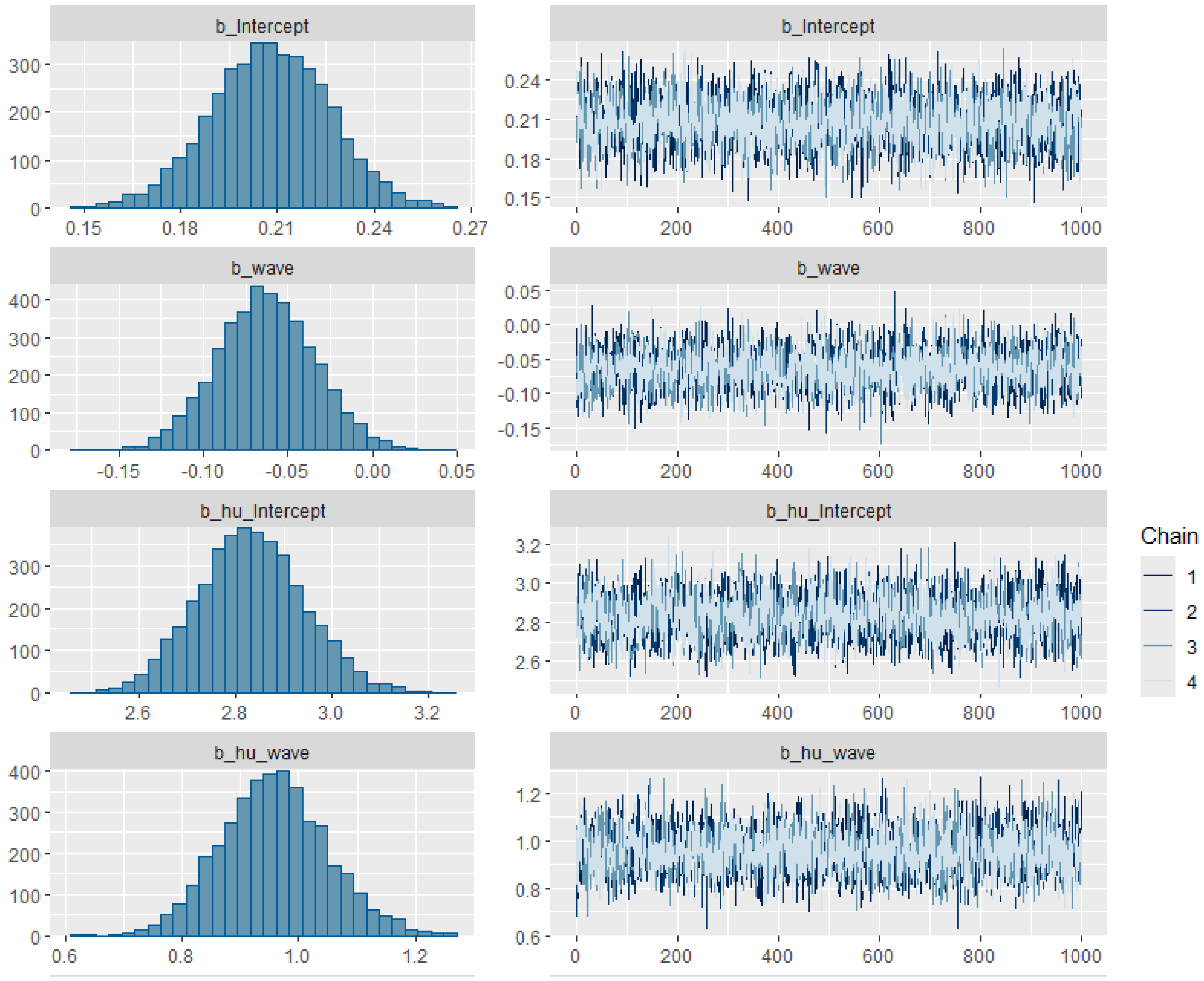

Figure S4. Trace plots for the unconditional A&E attendance model (fixed part)

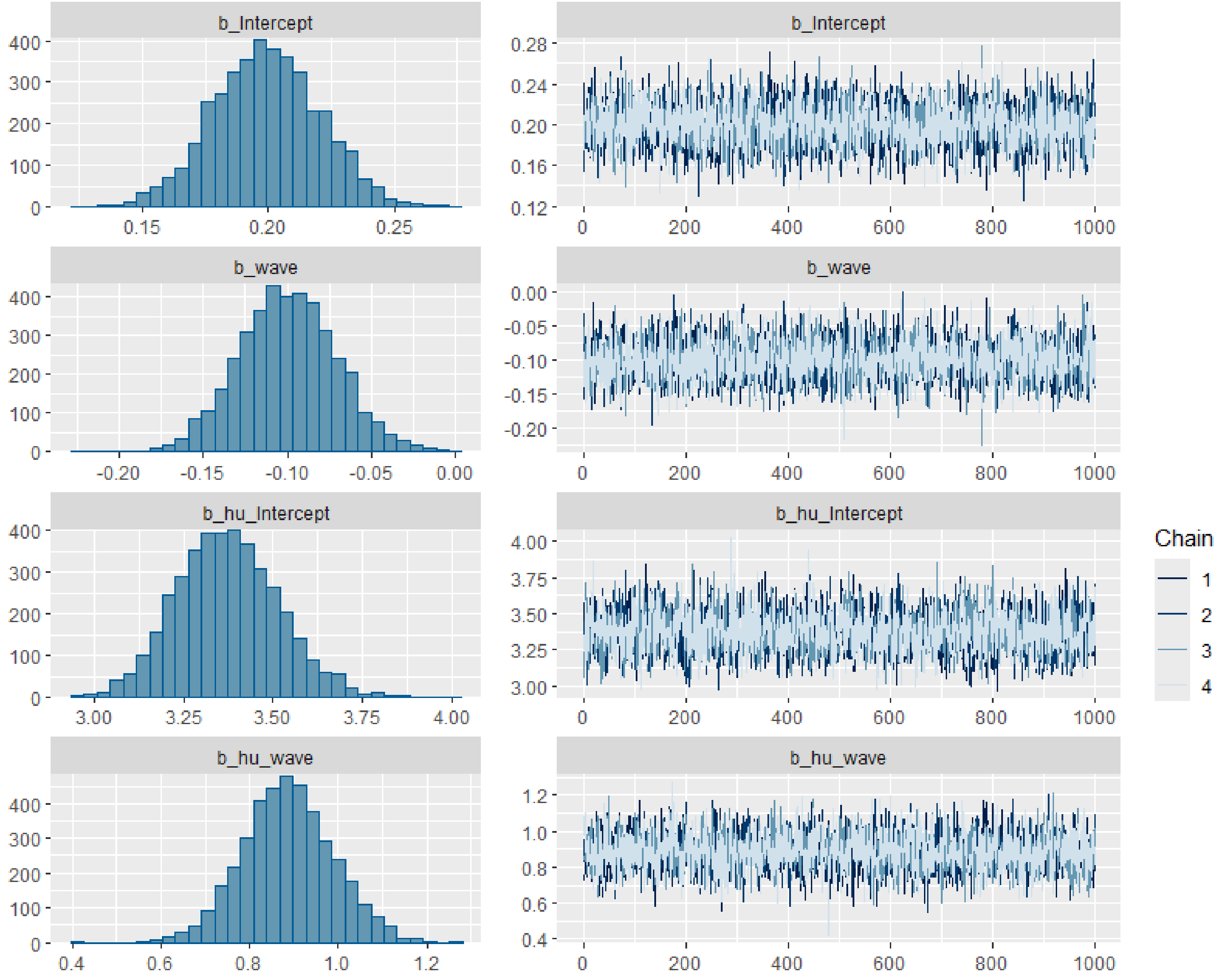

Figure S5. Trace plots for the unconditional hospital admission model (fixed part)
